# Vector control decision-making processes: Perspectives of twelve national malaria programmes across Africa

**DOI:** 10.64898/2026.05.12.26352987

**Authors:** Mercy Opiyo, Samuel Kweku Oppong, Elodie Vajda, Neil F. Lobo, Allison Tatarsky, Edward Thomsen

## Abstract

**Background:** Vector control is essential to malaria control and elimination. National Malaria Programmes (NMPs) must make complicated decisions about vector control in the face of evolving epidemiology, biological threats like insecticide resistance, a growing vector control toolbox, and an increasingly constrained funding landscape. The WHO recently published a manual on subnational tailoring of malaria strategies, but limited efforts have been made to understand how NMPs prioritize data and factors that impact decision-making in practice. This study explores vector control decision-making processes, enablers, and barriers across 12 African malaria programmes.

**Methods:** We conducted semi-structured interviews with 13 NMP managers or designated representatives from 12 African countries. Interviews were conducted virtually via Zoom or in-person, audio-recorded, transcribed, and thematically analyzed using content analysis. Participants described the interventions in use, decision-making factors, stratification approaches, perspectives on new tools, and operational challenges.

**Results:** Insecticide-treated bed nets (ITNs) and indoor residual spraying (IRS) are the core interventions in all countries, with limited but growing use of larval source management, mainly larviciding. Vector control tool selection is driven by WHO guidance, resistance profiles and patterns, epidemiological trends, operational feasibility, and donor funding priorities. Sub-national stratification is widely applied; however, limited analytic and modeling capacity hinder consistent application. Gaps in entomological data result in incomplete data availability to guide stratification. New vector control tools were perceived as promising options, albeit constrained by cost, limited evidence, regulatory delays, and community acceptability. Funding emerged as the dominant driver of decisions, shaping intervention choices regardless of country preference. Participants emphasized substantial gaps in vector control protection related to residual transmission, outdoor biting, insecticide resistance, and unprotected populations living in temporary structures or associated with high-risk occupations.

**Conclusions:** Vector control decision-making among NMPs is shaped by an interplay of scientific evidence, operational realities, and external funding dynamics. Strengthening entomological surveillance, enhancing SNT analytic and model output interpretation capacity, securing sustainable financing, and improving community engagement are critical to advancing tailored deployment of tools. Decision-support frameworks that reflect the complexities facing NMPs may further enhance evidence-based, context-specific vector control planning.

## Introduction

Malaria is a major public health challenge in sub-Saharan Africa, which accounted for approximately 94% of global malaria cases and 95% of deaths in 2024 (1). Significant reductions in malaria deaths and cases observed between 2000 and 2025 were largely attributed to wide-scale use of vector control (VC) interventions, particularly insecticide-treated nets (ITNs) and indoor residual spraying (IRS) (2,3). However, progress has plateaued in many countries due to widespread insecticide resistance, evolutionary adaptations in mosquito vectors, changing vector compositions, climate variability, limited access to and uptake of interventions, increasing population mobility, reduced funding, and structural health system constraints (1,4–6). Additionally, the heterogeneous nature of malaria transmission at sub-national levels necessitates targeted, stratified deployment of interventions rather than uniform, nationwide blanket approaches (1,7,8). These evolving conditions highlight the need for adaptive, evidence-informed, and context-specific VC strategies.

To address these challenges, the World Health Organization (WHO) recommends integrated vector management (IVM), a framework prioritizing evidence-based decision-making, intersectoral collaboration, and tailored combinations of interventions appropriate to local conditions (9). A key component of IVM is sub-national tailoring (SNT), which calls for deploying VC tools based on local epidemiology, vector behaviour and population dynamics, and operational feasibility (9–11). The recently published WHO manual on SNT of malaria strategies and interventions provides operational guidance on conducting stratification, selecting and prioritizing interventions across transmission settings, aligning implementation with available resources, and institutionalizing adaptive management through routine data use and periodic review (10). In parallel, WHO’s Global Technical Strategy (GTS) for Malaria 2021–2030 identifies SNT as essential to accelerate progress, optimize impact, and sustain gains toward elimination (12). In this context, and with the formalization of the SNT guidance, it is even more important and timely to document the NMP decision-making processes around VC to enable sharing of best practices and identification of common challenges.

Despite this strong policy focus, visibility into how NMPs make VC decisions, how operational, financial, and political factors influence these decisions, and how these factors differ across NMPs remains limited. Most research focuses on the efficacy or impact of interventions rather than the institutional processes guiding their adoption. Studies in Kenya, Zambia, and Tanzania suggest that, while NMPs aim to follow IVM principles, decision-making often suffers from limited entomological data, insufficient modelling capacity, donor-driven priorities, and operational bottlenecks (8,13,14). Regional assessments by WHO and the RBM Partnership to End Malaria show persistent gaps in the availability and use of surveillance data needed to effectively implement or operationalize SNT (12,15). Additionally, decisions concerning newer or supplemental tools such as dual-AI nets, spatial emanators (SEs), larval source management (LSM) approaches, as well as tools still in development like gene drive technologies, face uncertainties related to cost, regulatory approval, operational feasibility, and community acceptance (16–18).

Understanding these decision-making processes is essential, as they influence how limited resources are allocated, which populations are prioritized, and how effectively programmes respond to emerging epidemiological threats. These insights are essential for strengthening policy guidance, designing practical decision-support systems, building capacity, and ensuring more efficient, impactful, and context-appropriate deployment of vector control tools.

This study, therefore, sought to examine VC decision-making processes across twelve African malaria programmes. Through interviews with NMP representatives, we explore (1) current vector control interventions; (2) criteria and processes guiding decision-making; (3) the implementation and operationalisation of SNT; and (4) perspectives on new tools and persistent gaps. By documenting programmatic realities and decision-making pathways, this work aims to inform the development of practical, feasible, and evidence-aligned decision-support frameworks for malaria vector control.

## Methods

### Study design

Eighteen NMP managers from eighteen countries representing different regions and transmission settings across eastern, southern, western, and central Africa were purposively selected and invited via email to participate. The objective was to obtain insights into country-level decision-making processes for vector control interventions across diverse malaria transmission settings and decision contexts. The invitation email included the participant informed consent form, which was provided in advance to allow for informed review before participation. Participants were given a choice to either verbally consent during the interviews or sign consent forms (S1 File).

A total of 13 NMP managers and representatives from 12 countries expressed interest and consented to participate, representing eastern, southern, western, and central Africa. In instances where NMP managers were unable to attend due to conflicting commitments or busy schedules, they nominated programme representatives who possessed in-depth knowledge of vector control decision-making processes at the national or sub-national level.

Following agreement to participate, interview dates and times were arranged via email, taking into consideration time zone differences and participants’ availability. Each participant received a Zoom meeting link, and reminder messages were sent by email and text two to three days prior to the scheduled interview. When unforeseen circumstances prevented participation, interviews were rescheduled to accommodate new availability.

### Data collection and analysis

The semi-structured interview guide was developed by the study lead MO in close consultation with other members of the research team (SKO, NFL, AT and ET). The guide underwent multiple rounds of review and refinement until consensus was reached on a final version that adequately captured the study’s objectives and thematic areas (S2 File).

At the start of each interview, participants were informed that their responses would remain confidential, anonymized, and accessible only to the study team. All interviews were conducted virtually via Zoom, except for one that was carried out in person due to the participant’s limited virtual availability. In total, 13 participants from 12 countries took part in the study. Each country was represented by one participant, except for one country where both the national programme manager and the head of monitoring and evaluation participated.

Interviews took place between February and October 2025 and were conducted in either English or French and lasted approximately 60 to 90 minutes. Prior to each session, verbal consent to record the interview was obtained from participants. All interviews were audio-recorded and supplemented by written notes to ensure completeness and accuracy of the data. Audio recordings were subsequently transcribed, and each transcript was cross-checked against the corresponding audio file for quality assurance. Interviews conducted in French were first transcribed in French and then translated into English by a bilingual member of the research team proficient in both French and English (EV). The transcripts were subjected to content analysis, thematically coded, and analyzed using Dedoose software (19). Key excerpts were reported verbatim.

### Ethical review

This study was reviewed and approved by the Institutional Review Board of the University of California, San Francisco (approval no: 24-43074). The participants gave either written or oral informed consent prior to participation.

## Results

### Summary of the study participants

A total of 13 participants were interviewed across 12 countries. Of these, 31% were female and 69% were male. The sample included six NMP managers, one deputy NMP manager, five heads of vector control and surveillance units, and one head of planning, monitoring, and evaluation research. Regional representation consisted of three participants from East Africa, three from Southern Africa, six from West Africa, and one from Central Africa.

### Vector control tools and selection criteria

#### Vector control tools implemented

Participants consistently reported that ITNs and IRS remain the primary vector control interventions across their countries. ITNs were highlighted as the most widely implemented intervention and were frequently cited as the primary national VC strategy. Participants indicated that ITNs are commonly distributed through mass distribution campaigns, routine health services, and, in some countries, through additional channels such as schools. IRS was cited as the main strategy in two countries, where it is typically deployed in a targeted manner, either through focal or blanket spraying guided by localized malaria transmission patterns and operational feasibility. Seven countries conduct larviciding on a limited or pilot scale, two of which also implement targeted habitat modification. They emphasized that LSM has not yet been widely integrated into national VC programs, primarily due to financial constraints and limited operational capacity, and the need for stronger evidence on programmatic feasibility at scale.

#### Reasons and criteria identified for selecting vector control tools

The participants highlighted that the choice of VC tools is informed by a set of factors that include WHO guidance and recommendations, national strategic guidance, operational feasibility, and funding. In all countries, the main reason for selecting specific tools is their demonstrated effectiveness in local epidemiological and entomological contexts, especially in settings with increasing insecticide resistance. Insecticide resistance data was cited to play an essential role in these decisions, with resistance profiles directly informing the shift to next-generation ITNs such as dual-AI nets, PBO nets or the deployment of IRS in high-transmission areas.

> *“So, I think IRS and ITNs has served us very well. We have been able to gain some successes and reduced malaria incidence, especially in the areas with IRS but, If you look at the burden now and especially when we change insecticides, I cannot say that it is due to that, but what we began to see around that time, we see that the drop in cases is plateauing. And so, IRS is very effective and ideally if we had our own ways, we would be able to do it in all the areas that we’ve earmarked and even more if the funding allows us. But IRS tends to be quite expensive. And so, we are not able to do that. ITNs, on the other hand, are effective, but there’s always a catch of are people using it enough? Is it yielding the results because people are sleeping in it?” (Participant 4)*
>
> *“We conduct mosquito resistance surveys. We have entomological sentinel sites nationwide where we collect data on insecticide resistance and where we see that we need to deploy bed nets, we recommend specific types of net that are still effective on existing mosquito species. Where we have the high burden of malaria like high incidence with the documented resistance to pyrethroids, we recommend IRS as an effective intervention to be deployed.” (Participant 7)*
>
> *“Generally speaking, when it comes to mosquito nets, we use a systematic approach. It’s the insecticide that varies. Now, if resistance is proven through the studies that are carried out, or if the beginnings of resistance are reported by the entomologists who carry out the studies, we automatically ask the donors to tell us what we want. And recently, as I told you, on the basis of insecticide resistance monitoring, we wanted to leave pyrethroids behind and switch to double-impregnated ITNs with PBOs, which are currently used here. We started with the southern zone, where resistance was much stronger. And it was on that basis that we made our choice.” (Participant 11)*

The WHO guidance and the national malaria strategic plans (NSPs) were consistently mentioned as key documents that outline which interventions should be used and where, often based on burden stratification and sub-national epidemiological patterns.

> *“Tools are selected based on the national strategic plan* (*2024–2029*) *which highlights the tools to be used for vector control tools.” (Participant 1)*
>
> *“Results and budget also play a role. The ITNs and IRS are implemented because they are the ones available and there is strong recommendation from WHO.” (Participant 3)*
>
> *“So, when we had these new generation nets that have the potential to counteract the resistance for the vector that was resistant to pyrethroid only. Then in line with the WHO elimination framework, it [WHO elimination] guides that where the burden is highest, it’s always prudent to always deploy other high impact interventions. One of those high impact interventions though is costly, IRS is deployed in the highest burden districts to be able to bring it down to such levels, then we continue with the deployment of the ITNs.” (Participant 6)*

Yet financial limitations remained a recurring theme, often constraining the ability to implement IRS broadly, pushing programs to rely on cost-effective ITNs to maximize the number of people protected. The universal distribution of nets was widely embraced due to their affordability, safety, and protective value, reinforcing their place as a core intervention, particularly in the absence of newer, scalable tools.

> *“The threshold of selecting a vector control [tool], for ITNs, the intervention is determined by population demand. The criteria is that ITNs deteriorate, and every three years nets are distributed to the communities, and it’s [nets] given to the entire community. Quantification is done to determine demand. For mothers, the country uses WHO threshold, proportion of pregnant mothers within the population.” (Participant 1)*
>
> *“There are not enough funds to cover all high parasitemia sites according to the country’s recommendations, so those sites are covered with ITNs that are potent (new nets, IG2 first, PBO ITNs then standard ITNs).” (Participant 4)*
>
> *IRS is expensive but because of the dengue outbreak, [we] had to use IRS, even though wasn’t [sic] initially planned.” (Participant 8)*

The donor funding priorities were also highlighted as influential, as countries often align their choices with tools that external partners are willing to finance. However, discrepancies between donor priorities and country-defined needs frequently result in misalignment.

> *“They are [IRS and ITNs] the priority for the donor, so the country implements ITNs and IRS.” (Participant 3).*

### Vector control implementation strategy and distribution platforms

Participants described a comprehensive and multi-layered approach to implementing insecticide-treated nets (ITNs), reflecting a blend of centralized planning with diverse, context-specific delivery strategies. It was evident from the participants feedback that 1) every country’s implementation strategy focused on universal coverage which is defined by WHO as “ensuring all people having access to the full range of quality health services they need, when and where they need them, without financial hardship” and 2) to achieve universal coverage, countries implemented diverse distribution strategies i) mass campaign through door to door, and central distribution points, ii) continuous distribution through ANC, and iii) targeted distribution e.g. schools, specific high risk populations (HRPs), refugee camps and prisons.+

> *“So, for the mass ITN campaign, it [ITNs] starts first centrally in what we call the macro planning. So, we do the macro planning based on the population as indicated area we use the 1.8 people per one net. So, we get our population for that year when we want to distribute the ITNs divided by the 1.8, it will show us at least the number of nets then to […] data for n plus /minus, we add 10% buffer so […] in case we do the actual household registration, we have more households to receive nets […], we are at least covered. So, we do the macro planning based on the population and then after the macro planning [.]” (Participant 6)*
>
> *“For nets, the procurement is organized by the central level but based on the needs assessment conducted by health centers and in each health center catchment area, we have community health workers who conduct this needs assessment. They count the number of people who are eligible [and] the number of sleeping spaces per household.” (Participant 7)*

There is a wide range of distribution platforms with the aim of ensuring year-round access to vector control tools, especially for pregnant women and young children. Continuous distributions through health facilities (HFs), particularly antenatal care clinics (ANCs) and child welfare clinics (CWCs) were highlighted as a core channel. Two participants noted that school-based distribution, especially in boarding schools, has been piloted and proven effective, with plans to scale up once funding allows. Community-based strategies also play an important role, with local organizations and health workers facilitating door-to-door and village-level distributions, incorporating education and follow-up visits to ensure usage. Additionally, one participant pointed out that special populations, such as prisoners and refugees, are targeted through coordination with relevant institutions.

> *“For prison, we work with the Correctional Service Health department. They [Correctional Service Health department] also send us requests for bed nets based on the sleeping spaces and different prisons.” (Participant 7)*

Though the methods used in the distributions vary, the planning and procurement processes remain centralized, with national-level coordination based on population needs, household sleeping spaces, and buffer stock estimates. This centralized model aims to ensure consistency in coverage, while localized delivery mechanisms aim to improve accessibility, equity, and community ownership of ITN use.

### Malaria transmission context and stratification factors

Countries described that malaria stratification is based on categorizing risk levels across different geographic areas, primarily based on epidemiological data. Rather than applying a uniform national strategy, countries tailor interventions based on distinct risk strata, which include very low, low, moderate, high, and in some cases, very high burden. These strata classifications are often driven by malaria incidence data, with specific thresholds i.e., cases per 1,000 (e.g. 0–1 case, or 50+ cases per 1,000) used to define each stratum. Participants also highlighted that stratification enables more targeted decision-making, particularly by identifying districts or zones with high malaria transmission. For instance, one participant mentioned that they use ward-level incidence to classify areas, while others highlighted that they divide the country into zones according to both incidence and ecological transmission patterns. These zones often align with climatic conditions, such as rainy season transmission in Sahelian zones and areas with transmission throughout the year in more tropical regions. However, participants noted that, despite the availability of WHO guidance, many countries adopt stratification frameworks that remain imperfect due to challenges including accessibility constraints, limited capacity, and funding gaps. This approach allows programs to be more responsive, focusing resources where they are most needed and designing interventions that match the specific transmission intensity and context of each area.

### Role of new vector control tools in combating malaria transmission

#### Perspectives on current vector control tools and their adequacy for malaria programs

Overall, most participants believed that the vector control tools they are currently implementing are still working effectively. However, they highlighted the importance of first addressing key challenges that affect implementation, such as limited funding leading to reduced scale of implementation and inconsistent use (of ITNs). Further, participants felt that these issues should be resolved before considering the introduction of new tools while at the same time acknowledging that the current tools are mainly used indoors and are commodity-based.

> *“So, I think IRS and ITNs has served very well […] but there’s always a catch of are people using it enough? Is it [ITN] yielding the results because people are sleeping in it [ITN]?” (Participant 4) “[…] with tools we already have, these are insufficiently used. The problem starts here. I think LLINs are insufficiently used, even though these interventions work. We have interventions that work but are under-used.” (Participant 8)*
>
> *“Yeah, so in line with our strategic plan, the vector control tools that we have are adequate. The only challenge comes in when it comes to scale of deployment of the tools that we have.” (Participant 6)*

This leaves important gaps, especially with changing mosquito behavior, such as increased outdoor biting, the widespread emergence of insecticide resistance among key malaria vectors, and the difficulty of using existing tools in certain environments, as well as heterogenous human behavior impacting use or non-use of tools.

> *“I think these tools [ITNs and IRS] and the tools in the pipeline [e.g., spatial repellents] are sufficient. The real challenge is not so much in the tools but in the human behaviors that need to change, if buy-in isn’t good, the tool won’t be effective.” (Participant 10)*

The participants highlighted that considering such challenges, there is a need for complementary methods like larval source management (LSM), including larviciding, and possibly other tools such as spatial emanators (SEs) or housing improvements, beyond just relying on ITNs and IRS.

> *“This is what governments need to understand: firstly, malaria in Africa cannot be eliminated until the environment and sanitation are guaranteed.” (Participant 11)*
>
> *“The ITNs work well for the first 2 years and after that it [ITNs] does not work well. IRS works for 6 months, so its temporary. There is need for more like adding larviciding. There is need for new vector control tools, such as those that target the parasite and people. Tools for outdoor biting is needed.” (Participant 3)*

### Consideration when selecting new vector control tools

Participants reflected on a wide range of thoughtful and practical considerations that shape how countries decide which new vector control tools to adopt. A strong theme was community acceptability; tools must not only work technically but also be welcomed by the people expected to use them in their daily lives. Participants spoke about the need to truly listen to communities, ensure tools are easy to use, and make them feel part of the decision-making process.

> *“The community complains that they are not listened to enough - understand community perceptions of these tools to protect against malaria.” (Participant 10)*
>
> *“[…] even from the district’s level, to get people to use it [new tools], it has to be something that people accept [and] can easily use.” (Participant 4)*

Affordability and access were also central concerns, particularly in contexts where most funding comes from external donors. Tools that are cost-effective and able to reach many people were seen as more viable.

> *“Then the other thing is the cost. What’s the cost for that new tool? Just for us, our target is to ensure that too. Accessible and available or deployed to protect as many people as possible. So, if the cost is reasonable, then we know that [we can] protect many people.” (Participant 6)*
>
> *“The cost of the intervention is also important to consider: 70% of vector control funding comes from external resources.” (Participant 10)*
>
> *“The tools should be affordable to the country. The potential limitation of the tools includes the costs, efficacy of the new tools to address the resistance i.e. the resistance is a problem, the hope is the tools will be able to address the resistance challenge for a long period of time.” (Participant 3)*

Beyond cost, participants emphasized the importance of demonstrated impact on the disease, with a clear expectation that new tools should move countries closer to malaria elimination rather than being introduced for novelty’s sake.

> *“New vector control [tools] would be meant to address gaps in current vector control. But should not be deployed at a large scale, maybe subnational tailored intervention. And as we understand [new tools should be] […] at a subnational level and […] targeting of special areas. For repellent product, they should be targeting mainly hotels or boarding schools or prisons. Those groups [people] who may be spending the night outside like security staff.” (Participant 7)*
>
> *“Some of the key consideration of tool selection is impact and will take the country to the goal of elimination and not for academic purposes and publications.” (Participant 2)*

The context of implementation mattered deeply; participants spoke about tailoring tools to fit local transmission patterns, geography, and specific population needs rather than rolling them out uniformly. Entomological data, like insecticide resistance and mosquito behavior, was consistently mentioned as an essential input for making informed decisions about tool suitability.

> *“The IRS is selected based on the behavior of the vector which bite and rest indoors […]. IRS was able to target the vector and IRS is the main cornerstone of the country and has been for a long time.” (Participant 2)*
>
> *“When selecting new vector control [tools], the country needs to use entomology data to do analysis, consider the place where the tool is to be implemented and the incidence still remains high.” (Participant 3)*
>
> *“The prioritization of deployment is based on which tool is the most effective, and which will target the vectors in the most effective way taken into account their behaviors.” (Participant 4)*

Lastly, the technical capacity to manage and sustain new tools, especially complex ones like gene drive, was seen as a vital factor, with questions raised about whether local systems and staff are equipped to handle them responsibly. Altogether, these insights reflect a grounded, community-aware, and data-informed approach to evaluating new interventions, with a clear focus on equity, practicality, and local relevance.

> *“Do we have what it takes for us as a country to be able to implement that intervention […] the gene drive. Do we have the skill and what happens in case the mosquito continues with its mutation [.]. Are we going to be able to manage and so on, so those are some of the things that we put into consideration when considering the new tools.” (Participant 6)*

### Potential limitations of new vector control tools

Participants recognized the potential added value of introducing new or supplementary vector control tools; however, they also identified several significant barriers to their adoption and scale-up. A commonly cited challenge was limited tool availability, including lack of access by consumers, and delays in national regulatory approval processes even in cases where tools had already received WHO endorsement.

> *“[…] when it comes to them [new tools] being available in the local market, you find that they [new tools] are not there [….]. It [availability in local markets] limits people in benefiting from such new tools. By the community”. (Participant 6)*
>
> *“Challenges such as approval process of a new tool within the country can take a long a time for example, malaria vaccine has been approved by WHO but unfortunately the country has not approved the malaria vaccine for use. It takes time until its approved, and by the time its approved, the tool is perhaps not as good as the new tool in the market.” (Participant 1)*

Resource constraints were frequently emphasized, particularly the substantial upfront costs associated with many newer tools and the limited funding available through national budgets. Participants noted that donor reluctance to finance tools that are not yet fully recommended by WHO further restricts countries’ ability to diversify their vector control portfolios. WHO recommendations rely on evidence of public health value, which is often unavailable or slow to be produced.

> *“[…] even those available [tools] are not funded by the main funders of the programs and are not strongly recommended by WHO […]. I think there’s a need for policy change or guideline change. To make sure that where these specific tools are required, they [new tools] have a strong recommendation from WHO. So, this can back or support programs or countries in allocating funds, either from the government or from the main funders [such as] Global Fund, USAID, and many others. But if we see them [new tools] in the guidelines, they [new tools] are not really strongly recommended. Lack of evidence, of course. If you don’t have evidence, it is not easy to also get strong recommendation and also the buy-in by the funders.” (Participant 7)*
>
> *“Some potential limitation of the new tools will be on funding since some donors are not open to funding anything not approved by WHO and WHO take ages to approve a tool and its usually based on trials etc. For the tools that are WHO approved, the limitation will be acceptance by the people e.g. LSM could be acceptance, and this can be mitigated through engaging the community. Acceptance and proper use of the tool is a major limitation.” (Participant 2)*

Concerns related to tool performance also emerged. Some participants expressed uncertainty about the effectiveness of newer tools in addressing insecticide resistance or their durability across different ecological and seasonal conditions within their specific contexts.

> *“Efficacy of the new tools to address the resistance i.e. the resistance is a problem, we hope the tools will be able to address the resistance [which] has been a challenge for a long period of time”. (Participant 3)*

Participants highlighted that new vector control tools could play a role in malaria control; however, challenges such as community acceptance and user adherence were described as additional barriers that require intensive behavior change communication efforts, especially for new tools like gene drive. Similar concerns were raised by participants regarding ITNs, which also heavily relies on user adherence and intensive behavior change.

> *“[…] the ethical issues for example gene drive, so it has a limitation because it requires a lot of SBCC work for people to understand why you’ll be releasing the mosquitoes in their community”. (Participant 6)*
>
> *“The new tools can play a role in malaria control. But the biggest task is to change the behavior of those who use them.” (Participant 1)*

Finally, participants highlighted persistent gaps in local capacity, including reliance on external technical support and limited in-country expertise to guide implementation, monitoring, and evaluation. Overall, these perspectives reflect the financial, operational, and social considerations that shape the feasibility of adopting new vector control tools within malaria programmes.

> *“Capacitating individual researchers in the country rather than depending on outside capacity and tools developed from outside of Africa.” (Participant 1)*

### Gaps to address with new vector control tools

Participants identified several critical gaps that new vector control tools could help address. One of the most pressing issues is the changing behavior of malaria vectors, such as biting outdoors, earlier in the evening, and continuing into the morning, making current indoor-focused tools like ITNs and IRS less effective.

> *“They [mosquitoes] are now biting outside. And they [mosquitoes] are biting earlier than before. We can also have those [mosquitoes] feeding on both animals and human which is also another challenge and they’re [mosquitoes] creating a gap in existing interventions.” (Participant 7)*
>
> *“Based on ento [sic] data, mosquitoes bite until 8-10 am, when people are out of bednets.” (Participant 10)*
>
> *“And we also realized that LLINs, of course, protect indoors when people are asleep, but when they’re [people] not asleep and there’s activity, it’s not always possible. So, we don’t have any other tools available.” (Participant 9)*

Participants also highlighted that some areas remain entirely unprotected due to limitations in existing interventions or funding gaps. For example, two participants highlighted that people living in temporary or unsprayable structures are excluded from indoor spraying programs, and some communities still rely on traditional methods like herbs due to lack of access.

> *“I saw there’s specific gaps that they [new tools] can help in addressing. Let me pick IRS, for example. So, for IRS, we have what we call eligibility criteria. So, the criteria entail […], the structure before it is sprayed, it must be a permanent building, not temporary, so that structure will be sprayable surfaces which should be made of materials that are either permanent or semi-permanent. Then people should be sleeping in that structure at least not less than six months. So, for us we consider the new tools that they [new tools] can be able to address some of those issues, for example, where the structure is not eligible. You cannot spray it, but you still need to protect those people […] [from] mosquito bites. So, if there’s a way of having other tools, those tools can help you in protecting those people that are not eligible to receive the other [IRS] intervention because of the structure that they [people] live in. So, for me, it can help with me [sic] ensuring that their [new tools] protection to the people that we are targeting.” (Participant 6)*
>
> *“New tools can address different gaps […] like urban malaria transmission in urban settings [where] tools like IRS are hard to implement. For example, urban malaria transmission in [some towns], IRS cannot be conducted due to various challenges like structures; construction of the buildings and various factors and other tools can come and fill in this gap. We have succeeded due to IRS and ITNs, but some towns remain problematic and even ITNs are not well accepted (low acceptance) and so new tools can be used in such settings.” (Participant 1)*

Low community acceptance of in some areas of current tools like ITNs or IRS, further limits their impact. Participants also drew attention to new and expanding mosquito larval sites caused by development projects like mining or agriculture, which current interventions don’t adequately cover

> *“There are also risks linked to activities (mining, forestry) where mosquito nets are not easily used. We need to identify the gaps in protection and identify the most vulnerable populations.” (Participant 10)*
>
> *“We have now new breeding [larval sites] sites emerging probably due to some […] economic projects like rice field, like mining activities, water dams. And this is contributing to mosquito breeding [larval] site that cannot [sic] address by existing indoor interventions.” (Participant 7)*

### Drivers of decision making in vector control

Participants identified a range of interrelated drivers of decision-making in vector control, with resource availability as a key influence. Funding constraints, particularly those tied to donor priorities, often dictate what interventions can be implemented, regardless of local preferences or needs.

> *“Oh, so, a typical case is the fact that Global Fund does not support larviciding for now. And so, it means that in a funding request, in our proposals, there’s no way we can include larviciding, though it’s something that we probably would have wanted to scale up throughout the country. So that is one example. And so even the decision of what you can do or what you can implement is largely based on what’s [sic] the funding supports. And that said, once you have a number that the funding arrangement supports, now the question is how these funders affect the decisions you make. Usually for Global Fund and even PMI, we are able to send the request based on a strategic plan as best as it fits within the funding model of the funding agency.” (Participant 4)*
>
> *“Cost parameters are considered if it fits with the specification and quality of the products. In terms of funding, we abide to the ceiling of what is available according to donors. Donors like PMI adjust and shape priorities.” (Participant 1)*
>
> *“Donors have a very big role - influence selection. Misalignment between the country’s vision has for vector control in their country and what the donors have in mind.” (Participant 10)*

Participants emphasized that cost, population protected, and procurement challenges heavily influence tool selection.

> *“Then the cost [of] that intervention […] also influences how much it costs to protect a certain population. If it’s [to] be very expensive just protecting a small population […] certainly we may not consider that. [Cost] influences on us picking it [intervention]. Then […] also the coverage of certain intervention [interventions]. How many people [we] would be able to protect when we have deployed it [intervention] […] so that also influence [sic]. For example, if [we] were to put IRS and ITNs, if they [we] had to pick say which one would cover more people at a minimal cost at one time.” (Participant 6)*

Participants reported that key considerations in selecting vector control tools included the overall cost of protecting populations—often framed in terms of the number of people protected by a given tool—and the perceived effectiveness of the intervention. However, cost-effectiveness, defined as the cost per case or death averted, was not explicitly mentioned. This is notable given that cost-effectiveness is a central metric recommended by the WHO to inform public health decision-making and resource allocation.

Epidemiological data play a central role in targeting burden areas, guiding where interventions such as ITNs or IRS are prioritized. Participants also noted that decisions are influenced by the demonstrated impact of tools; when an intervention shows evidence of success, such as significant reductions in malaria cases, it is likely to be considered and scaled up.

> *“Epidemiology is crucial to guide decision as the goal is to reduce the case of malaria [sic], the trends are assessed and high burden areas are assessed and prioritized, decision is made based on that and Global Fund commitments.” (Participant 3)*
>
> *“Based on epidemiology, that’s how the country was able to reintroduce ITNs so it’s important for decision making and of other interventions.” (Participant 2)*

All participants also highlighted that entomological factors including resistance profiles and vector susceptibility, further inform choices around which tools are most appropriate in a given context.

Political dynamics were described in terms of their dual effect on decision making, sometimes enabling but often stalling progress due to inaction or misaligned priorities between national civil servants working on health and politicians. Political interference was reported also as an obstacle, particularly when appointments to key roles were influenced by favoritism rather than technical competence, undermining operational effectiveness. Social and cultural acceptance of interventions was also highlighted as essential, with community engagement considered key for success and in decision-making. Moreover, practical factors such as the availability of materials or products locally can influence whether an intervention is feasible. Overall, decision-making in vector control was described as a complex process, constantly negotiated and inextricably linked with scientific evidence, funding realities, political and social context.

> *“Politics has a certain influence that isn’t always positive, in the sense of inaction. We’ve had regions for 7 years that haven’t received any LLINs because the state took charge of them. Lack of prioritization on the part of politicians.” (Participant 10)*
>
> *As for political, at intervention level, politics does not really play a big role, for example the intervention to use, that is an internal decision the program can make. Political influence (decision) can play a role in cases where for example if the country wants larviciding as a potential tool and thinks it’s [larviciding] more effective than ITNs or IRS, some political influence will play a role.” (Participant 3)*

### Sub-national stratification and tailoring strategy

#### Presence of SNT and support systems

Participants shared a mixed picture regarding the presence and use of sub-national stratification and tailoring (SNT) and decision-support systems in vector control. Ten countries reported having SNT in place, often as part of strategic planning processes or midterm reviews, with stratification used to guide intervention targeting and tailoring.

> *“SNT planning is done annually. Depending on the data available. But what is more recommended is at the end of the year, with all the periods, because there are periods of high transmission because transmission in January is totally different from transmission in March”. (Participant 11)*
>
> *“SNT is done as part of the process of developing strategic plan and midterm review for the strategic plan.” (Participant 3)*

However, this practice was not uniformly adopted; two participants indicated that SNT had not been implemented in their contexts. All participants noted that formal SNT committees were largely absent. Most respondents indicated that stratification tasks were handled by M&E teams and supported by partners, rather than through dedicated technical committees. While some countries reported working with academic partners or international institutions to support modeling and decision-making, in-country modeling capacity was generally described as limited or small-scale. Modeling of SNT scenarios, where present, was often donor- or partner-driven rather than embedded within national malaria programs.

> *“Okay, so we don’t have say specific mathematical modelers. But we have the biostatistician who heads the unit for M & E.” (Participant 6)*
>
> *“Modelling capacity is not yet established.” (Participant 3)*

#### Roles of SNT partners

Participants described a wide range of institutional partners playing critical and complementary roles in SNT processes, each contributing according to their technical strengths. Key partners such as Clinton Health Access Initiative (CHAI), WHO, Malaria Atlas Project (MAP), Harvard University, Northwestern University, and Zambia Health Analytics (ZHA) were noted for their contributions to data analysis and modeling, providing essential insights for tailoring malaria interventions. These institutions often work collaboratively with NMPs to forecast intervention impacts, generate visualizations, and analyze complex datasets. WHO was described as a key player, offering technical guidance, capacity building, and support throughout the stratification process from data review to training and stratification. Participants also highlighted that partners, including CHAI and WHO, also contribute directly to malaria report development, ensuring evidence is synthesized for action. In terms of coordination, participants highlighted the role of group discussions involving all partners like CHAI, Global Fund, WHO, Malaria Consortium, and others to align strategies and decisions. Capacity building was also mentioned as an area of support, with institutions like African Leaders Malaria Alliance (ALMA), Gates Foundation (GF), and WHO helping to strengthen technical skills in modeling, data management, and monitoring. Lastly, partners such as PATH and GF were noted to support the infrastructure side of data management, hosting data repositories and supporting quality audits to ensure robust decision-making systems.

#### Reasons for conducting SNT

Participants highlighted various reasons for SNT, rooted in both practical and strategic considerations. A key driver was the desire to maximize impact by focusing interventions where they would yield the greatest benefit, especially in high-burden or at-risk areas.

> *“To focus the interventions where the greatest impact can be achieved.” (Participant 3)*

Given that much of the operational work and implementation takes place at the sub-national level, tailoring strategies to the specific epidemiological and contextual realities of those areas was seen as essential.

> “[…] most of the work happens at the subnational areas. For instance, here [specific areas], you can’t come and say, okay, I’m going to do nothing in this place. It won’t work.” (Participant 5)

Limited funding emerged as a critical factor, prompting programs to adopt more efficient and targeted approaches to ensure available resources were used judiciously. Additionally, monitoring malaria trends at finer geographic levels allowed for more nuanced decision-making, particularly in identifying low-transmission areas within larger districts or provinces.

#### Challenges of conducting SNT

Participants highlighted various challenges that limit the effective implementation of SNT for malaria vector control. A key barrier consistently noted was limited technical capacity, particularly in modeling, data analysis, and interpretation of modeling outputs. Many participants described a lack of trained personnel and basic understanding of SNT concepts, which hinders both the uptake and practical application of these tools.

> *“Understanding the modeling and the concept, expertise and basic knowledge is lacking.” (Participant 3)*
>
> *“Need for greater modeling capacity.” (Participant 10)*

This capacity gap was compounded by challenges with the availability and quality of entomological data. Gaps in entomological sampling across regions, poor data accuracy, and limited digitization were cited as critical issues that affect the reliability of using entomological data to guide stratification and decision-making.

> *“Challenge of entomological data availability. There are data gaps which cannot be collected throughout the country. This affects modeling if the quality and completeness of the data is not adequate.” (Participant 10)*
>
> *“Digitization is one of our major challenges. With the new technologies, it’s faster and the information goes directly onto the platform, which saves you a lot of time, and allows you to track the evolution of malaria over time, and […] to see the distribution of the territory.” (Participant 11)*

Furthermore, one participant pointed to difficulties in aligning partners around a common vision, given the diversity of expertise and institutional agendas, which can lead to fragmentation rather than coordinated action.

> *“Aligning the partners to the same goal as the partners have different expertise, so partners have to be kept aligned and on the same goal. There is some level of competition, and the program director always has to find the best way to harmonize and drive the key goal with all partners involved.” (Participant 3)*

A summary of all factors influencing vector control decision making is described in (Table1)

**Table 1.** Factors shaping vector control decision-making in ministries of health.

| Tier 1. Binding decision constraints |  |  |  |
| --- | --- | --- | --- |
| *Factors that effectively determines what is implemented, regardless of preference. These are not negotiable factors. If a tool fails here, it does not move forward, no matter how innovative or promising it is.* |  |  |  |
| # | Factor | How it influences decisions | Evidence from interviews |
| 1 | Funding availability and donor priorities | This sets the ceiling for what is feasible and within the funding model. Misalignment between locally preferred tools and those prioritized by donors. Tools not supported by major donors are excluded, even when they are technically preferred. | Funding availability emerged as the dominant driver. Lack of donor support constrained larviciding, while IRS remained limited despite its perceived impact and supporting evidence. |
| 2 | WHO recommendations and guideline strength | WHO acts as the policy authority. Strong WHO recommendations facilitate donor support and national level adoption. | WHO guidance cited as foundational; weak, conditional, or supplemental recommendations limit uptake of new tools. |
| 3 | Unit cost and population coverage | Favors interventions that protect the greatest number of people at the lowest unit cost, often prioritizing ITNs. | Decisions framed around people protected per cost; IRS constrained by expense despite perceived effectiveness. |
| 4 | National strategic plan (NSP) alignment | Determines what is administratively and politically defensible in national planning and procurement cycles. | Tool selection repeatedly tied to National Malaria Strategic Plans and planning cycles which outlines both the choice of interventions and their geographic deployment. |
| **Tier 2. Evidence-driven technical modifiers** |  |  |  |
| *Factors that shape where, how, and which version of a tool is used. These factors matter a lot, but they only operate within the boundaries set by Tier 1. These factors optimize decisions rather than define them.* |  |  |  |
| 5 | Epidemiology burden and trends | Determines prioritization of areas and intensity/type of intervention deployment. | Incidence thresholds used for stratification; high-burden areas prioritized for IRS or next-generation nets. |
| 6 | Insecticide resistance (profiles) | Guides selection of ITN type and IRS insecticide choices. | Resistance data drove shifts from pyrethroid only nets to PBO and dual-active ingredient nets, and informed IRS targeting. |
| 7 | Entomological surveillance data quality | Enables or limits confident tailoring; gaps reduce precision of decisions. | Data gaps and incomplete coverage weaken stratification and modeling. |
| 8 | Sub-national tailoring capacity | Allows targeted deployment rather than blanket approaches; limited capacity reduces consistent application. | SNT widely used but inconsistently applied due to limited analytic and modeling capacity. |
| 9 | Demonstrated impact of tools | Supports continuation or scale-up when reductions in cases are observed. | Interventions showing evidence of success were described as more likely to be scaled. |
| **Tier 3. Contextual and implementation moderators** |  |  |  |
| Factors that influence feasibility, sustainability, and real-world effectiveness. These factors rarely decide what tool is chosen, but they strongly determine effectiveness/efficacy. |  |  |  |
| 10 | Community acceptability and behavior | Influences uptake, usage and sustained effectiveness; low community buy-in limits impact. | Participants emphasized that tools fail without community buy-in; behavior change repeatedly cited as critical for both current and new tools. |
| 11 | Operational feasibility | Determines deliverability and sustainability; logistical constraints limit scale. | IRS constrained by eligibility of structures; LSM limited by operational capacity. |
| 12 | Regulatory approval timelines | Can delay or block adoption, even after international endorsement. | National approval processes described as slow, with delays noted even after WHO approval. |
| 13 | In-country technical capacity | Capacity impacts readiness to implement complex tools and sustain monitoring and evaluation. | Concerns raised about implementing and managing complex tools such as gene drive. |
| 14 | Political dynamics | May enable or stall implementation through prioritization, inaction, or interference. | Political inaction and misalignment described as barriers to distribution and implementation. |
| 15 | Partner coordination | Affects coherence of planning and execution; misalignment between partners may fragment strategies. | Competition, differences in expertise and challenges in aligning partners to common goals were described during SNT processes. |

## Discussion

This multi-country qualitative assessment provides important insights into how NMPs navigate the increasingly complex landscape of malaria vector control. While most peer-reviewed literature evaluates the efficacy and effectiveness of interventions, fewer studies explore how the vector control decisions are made at national level. These findings underscore that vector control decisions in African NMPs are influenced by an interplay of epidemiological evidence, institutional processes, donor dynamics, operational and financial constraints, and the emerging need for sub-national tailoring (SNT).

NMPs consistently emphasized the importance of WHO recommendations, entomological surveillance, and epidemiological burden in guiding vector control choices. This aligns with IVM principles and global guidance (9,12). Resistance data were particularly influential, driving the transition from pyrethroid United States Agency of International Development (USAID) and the President’s Malaria Initiative (PMI) was being dismantled, making funding constraints even more challenging. Donor priorities, especially those of the Global Fund and PMI, strongly influenced which interventions could be implemented, often resulting in the prioritisation of ITNs due to their lower unit cost and broad protection potential (number of people protected). Similar observations have been reported across Africa and other regions, where dependence on external funding limits national flexibility and long-term planning for more expensive interventions such as IRS (20,21).

Most countries reported having SNT processes in place, yet the degree of implementation varied widely. Weak entomological surveillance systems and fragmented data, insufficient modelling and model interpretation capacity, and limited human resources hindered effective use of stratification and SNT in planning. These challenges are consistent with regional assessments that highlight persistent gaps in entomological data quality and SNT capacity across the continent (14,22). Moreover, while SNT is increasingly recognized as essential for targeting interventions, our findings indicate that many NMPs rely heavily on partners for data analysis and modelling. Such reliance risks undermining sustainability and national ownership and may delay or complicate decision-making.

Although participants emphasized that existing vector control tools are largely sufficient and that the priority is to address implementation gaps associated with their deployment (like usage and coverage), they also expressed strong interest in new vector control tools. There was interest in tools that target gaps related to outdoor and early-evening biting, unsprayable structures, and high-risk occupational groups with tools like larviciding consistently highlighted as one of such tools that can mitigate these gaps. These gaps are well documented in the literature, with residual transmission increasingly recognized as a major barrier to achieving malaria elimination (23–25).

However, enthusiasm was attenuated by concerns regarding cost, regulatory delays, limited in-country evidence, acceptability, technical capacity for evaluation and analysis and operational feasibility. These concerns reflect broader debates around the adoption of new tools, where innovation outpaces the operational capacities and regulatory systems of many malaria-endemic countries (16–18). Tools such as spatial emanators and gene drive were viewed as promising but requiring stringent evidence, community engagement, and substantial capacity-building before widespread adoption. Notably, the emphasis on community acceptability aligns with growing recognition that behavioural factors and social norms strongly influence vector control uptake and effectiveness (18,26–28).

The study highlights several operational realities that influence vector control decision-making. These include inadequate and often unstable funding, with a strong reliance on external donors; limited in-country entomological and modelling capacity to inform evidence-based planning; and misalignment between policy processes and programme priorities. Additional challenges include persistent gaps in protecting populations residing in unsprayable or temporary structures, as well as the emergence of new mosquito larval sites associated with infrastructure development and other large-scale development projects. These challenges are frequently cited across African malaria programmes and require multi-sectoral action (14,20,21,29,30).

This study has several limitations. First, the qualitative design relies on self-reported perceptions, which may be subject to recall bias and social desirability bias (31,32). Second, although the sample was geographically diverse, it included only 12 national malaria programmes and therefore may not fully capture the breadth of vector control decision-making contexts across Africa. Finally, the study did not include direct observation of decision-making processes and instead relied on participant accounts, which may limit insight into real-time practices.

## Conclusion

Vector control decision-making in African malaria programmes is a complex process shaped by normative guidance, scientific evidence, resource constraints, operational realities, community needs and donor dynamics. While NMPs aim to ground decisions in entomological data and insecticide resistance profiles, epidemiological data, and WHO guidance, limited funding and technical and operational capacity often determine what is feasible in practice. Strengthening entomological surveillance systems, investing in SNT and modelling capacity, improving coordination with partners, and expanding sustainable financing mechanisms are essential to support more tailored and effective deployment of vector control tools. As new interventions emerge, decision-support frameworks that explicitly incorporate use cases for the expanding vector control toolbox and considerations of real-world constraints will be critical in enabling NMPs to respond to evolving malaria transmission landscapes.

## Acknowledgement

We would like to express our appreciation to all National Malaria Programme (NMP) managers and country representatives across Africa who participated in the interviews. Chat-GPT (free online version: GPT-5.3 model) was used to improve sentence clarity during manuscript preparation in some specific places.

## Data availability statement

All data underlying the results presented in this study are included in the manuscript. Given the identifiability of participants, the full transcripts cannot be provided. Requests for data should be made to the University of California, San Francisco at. Extended supplementary files are also available in the repository without restriction.

Figshare: *Vector control decision-making processes Perspectives of twelve national malaria programmes across Africa* (https://doi.org/10.6084/m9.figshare.32108593)(33).

This project contains the following files:

- S1 File. Informed consent form Vector control decision-making processes Perspectives of twelve national malaria programmes across Africa (DOCX)
- S2 File. Key Informant interview guide Vector control decision-making processes Perspectives of twelve national malaria programmes across Africa (DOCX)

## Author contributions

Conceptualization: MO, AT, NFL, ET

Methodology: MO, SO, NFL, AT, ET

Data collection and curation: MO, EV, ET

Analysis: MO

Writing original draft: MO

Writing - review & editing: MO, SO, NFL, AT, ET

